# COVID-19 vaccine uptake among older people in relation to sociodemographic factors –cohort results from southern Sweden

**DOI:** 10.1101/2021.08.12.21261981

**Authors:** Malin Inghammar, Mahnaz Moghaddassi, Magnus Rasmussen, Ulf Malmqvist, Fredrik Kahn, Jonas Björk

**Affiliations:** Department of Clinical Sciences Lund, Section for Infection Medicine, Skåne University Hospital, Lund University, Lund, Sweden; Social Medicine and Global Health, Department of Clinical Sciences Malmö, Lund University, Malmö, Sweden; Clinical Studies Sweden, Forum South, Skåne University Hospital, Lund, Sweden; Division of Occupational and Environmental Medicine, Lund University, Lund, Sweden

**Keywords:** SARS-CoV2 vaccine uptake

## Abstract

The aim of this cohort study was to investigate sociodemographic determinants of COVID-19 vaccine uptake in the 70+ age group in Skåne county, Sweden (n = 216 243 at baseline). Uptake of the first dose was high (91.9%) overall, but markedly lower (75.3%) among persons born outside the Nordic countries. Vaccine uptake was generally satisfactory among native Swedes also in areas with lower socioeconomic status, but dropped substantially among non-Nordic born in those areas. The identified clusters of unvaccinated older people, mainly representing ethnic minorities in disadvantaged areas, warrants intensified efforts regarding tailored communication, easier vaccine access and local engagement.

**Key points:**

- COVID-19 vaccine uptake in the 70+ age group in Skåne county, Sweden, was high (91.9%) overall, but markedly lower (75.3%) within the group born outside the Nordic countries
- Inverse associations between indicators of neighbourhood deprivation and vaccine uptake were observed, which lowered the uptake among persons born outside the Nordic countries further
- The identified clusters of unvaccinated older people, mainly representing ethnic minorities in disadvantaged areas, warrants intensified efforts regarding tailored communication, easier vaccine access and local engagement

## Introduction

The emergence of more infectious severe acute respiratory syndrome coronavirus 2 (SARS-CoV-2) variants has stressed the importance of obtaining high and equally distributed vaccine uptake in the population to protect against coronavirus disease 2019 (COVID-19) [1, 2]. The COVID-19 vaccination has been a race against population protection with specific logistic challenges compared to the seasonal influenza vaccination programs, which are generally of proactive nature, focusing on targeted rather than general populations [2, 3].

Recent evidence from UK suggest lower COVID-19 vaccine uptake in deprived areas, especially among older persons [4]. Clustering of high-risk individuals with lower protection is worrisome [2]. The aim of the present study was to investigate sociodemographic determinants of COVID-19 vaccine uptake among older persons in a general population from Sweden.

## Methods

### Study cohort

The study cohort included all persons born 1951 or earlier (aged ≥70 years) during 2021, residing in Skåne county, Sweden, on 27 December 2020 (baseline) when vaccinations started (n = 216 243). The cohort was followed for 160 days, until June 4^th^ 2021. Individuals who died or moved out from the region during follow up were censored on the date of death or relocation, yielding 211 126 persons at the end of follow up.

Linkage from different data sources was facilitated using the personal identification number assigned to all Swedish citizens for life. Individual-level data on country of birth, civil status, residency and vital status were obtained from the Swedish Total Population Register. Data on vaccination date, type of vaccine and dose during follow up were obtained from the National Vaccination Register at Public Health Agency, Sweden. Area-level data on socioeconomic conditions at baseline were obtained for each postal code area from Statistics Sweden for the residential addresses. There are 1 353 postal code areas in Skåne county, with a mean total population of 1033 persons in each area.

### Statistical analysis

Statistical analyses were conducted in Stata SE 14.2 (Stata Corp.) and IBM SPSS Statistics 26 (SPSS Corp.). The study outcome was reception of at least one dose of any COVID-19 vaccine (Pfizer-BioNTech, AstraZeneca or Moderna) by the end of follow up, hereafter referred to as vaccine uptake. Survival curves based on the Kaplan-Meier method was used to investigate the time course of the vaccine uptake during follow up. Associations between sociodemographic factors and vaccination uptake were modelled with logistic regression and reported as odds ratios (ORs) together with 95% confidence intervals (CIs).

Vaccination with the second dose was still ongoing in the study population by the end of follow up, and differences in uptake across group could therefore not be assessed with certainty. However, time to reception of the second dose was investigated in relation to country of birth as a sensitivity analysis. This analysis was restricted to persons who received vaccine from Pfizer-BioNTech or Moderna since the dose interval for the vaccine from AstraZeneca was too long (10 weeks) to be included.

## Results

The overall vaccine uptake, i.e. the proportion of the 70+ population with at least one dose received, was high (91.9%) during the first 160 days of the vaccination program, with little variability across the 33 municipalities in the region (range 88.1 - 96.9%). Larger variability was, however, observed across the 1 290 postal code areas with a total population with ≥ 100 individuals, where the 2.5 – 97.5 percentile range for vaccine uptake was 72.9 – 98.8%. The Pfizer-BioNTech COVID-19 vaccine constituted 70.4%, AstraZeneca 24.9% and Moderna 4.8% of all first doses.

Relatively low vaccine willingness was observed among persons born outside the Nordic countries, representing 10% of the study population, with an uptake that seemed to plateau at 75% after 160 days of vaccinations (Figure S1A). However, there was considerable variation in vaccine uptake within this group. High uptake was observed among persons born in Western Europe, North America, Australia or New Zeeland (86%; n = 4 542), whereas lower uptake was noted among persons born in Africa (67%; n=635), Middle East (68%; n=2 732), Eastern Europe, former Yugoslavia, Russia or the former republics of the Soviet Union (73%; n=11 974), and Asia, Central- or Latin America (n=1 919; 77%; Table S1). Associations between sociodemographic factors and vaccine uptake generally appeared stronger among persons born outside the Nordic countries in terms of percentage point differences (Table 1). In the multivariable analysis for the total study population, living alone was clearly associated with lower vaccine uptake, whereas modest individual-level associations with sex (lower uptake among females) and age (higher uptake among 80-89 year olds) were observed (Table S1). Living in areas with high proportions of people born abroad, low educational level, high proportions of tenants, low income and low employment level, were all associated with lower vaccine uptake. The sensitivity analysis revealed no apparent differences in the uptake of the second dose in relation to country of birth (Figure S1B).

**Table 1.** COVID-19 vaccine uptake of the study population (n = 211 126 aged 70+) after 160 days of vaccinations, in relation to country of birth, individual- and area-level characteristics.

|  | Country of birth |  |  |  |
| --- | --- | --- | --- | --- |
|  | Nordic country |  | Other |  |
|  | N (%) | Uptake, % | N (%) | Uptake, % |
| **Total** | 189 152 (100) | 93.8 | 21 974 (100) | 75.3 |
| **Individual-level characteristics** |  |  |  |  |
| Age |  |  |  |  |
| 70-79 | 118 896 (62.9) | 93.5 | 14 665 (66.7) | 73.9 |
| 80-89 | 56 204 (29.7) | 94.5 | 6 177 (28.1) | 78.9 |
| 90+ | 14 052 (7.4) | 93.6 | 1 142 (5.2) | 74.5 |
| Sex |  |  |  |  |
| Females | 102 938 (54.4) | 93.6 | 11 871 (54.0) | 74.0 |
| Males | 86 214 (45.6) | 94.0 | 10 103 (46.0) | 77.0 |
| Civil status |  |  |  |  |
| Married | 98 274 (52.0) | 96.1 | 10 834 (49.3) | 79.1 |
| Widow/widower | 40 697 (21.5) | 94.4 | 4 722 (21.5) | 75.7 |
| Divorced | 33 657 (17.8) | 89.9 | 5 464 (24.9) | 68.6 |
| Single | 16 524 (8.7) | 86.4 | 954 (4.3) | 69.1 |
| **Area-level<sup>a</sup> characteristics** |  |  |  |  |
| Born abroad (%) |  |  |  |  |
| < 20 | 130 892 (69.2) | 94.5 | 7 185 (32.7) | 80.9 |
| 20 – 39 | 44 982 (23.8) | 93.0 | 7 305 (33.2) | 75.1 |
| 40 – 59 | 12 281 (6.5) | 89.7 | 5 878 (26.7) | 70.5 |
| ≥ 60 | 997 (0.5) | 85.0 | 1 606 (7.3) | 69.5 |
| Education, tertiary (%) |  |  |  |  |
| < 25 | 16 367 (8.7) | 91.1 | 3 020 (13.7) | 70.1 |
| 25 – 39 | 75 130 (39.7) | 93.0 | 9 261 (42.1) | 74.2 |
| 40 – 59 | 66 363 (35.1) | 94.6 | 6 221 (28.3) | 77.4 |
| ≥ 60 | 31 292 (16.5) | 95.4 | 3 472 (15.8) | 79.1 |
| Tenants (%) |  |  |  |  |
| < 10 | 80 054 (42.3) | 94.5 | 6 232 (28.4) | 79.9 |
| 10 – 49 | 70 426 (37.2) | 94.2 | 6 730 (30.6) | 77.6 |
| 50 – 74 | 26 688 (14.1) | 92.3 | 4 391 (20.0) | 73.5 |
| ≥ 75 | 11 984 (6.3) | 90.5 | 4 621 (21.0) | 67.6 |
| Yearly income, median (kSEK) |  |  |  |  |
| < 200 | 36 890 (19.5) | 91.3 | 10 113 (46.0) | 70.9 |
| 200 – 249 | 82 841 (43.8) | 93.7 | 7 106 (32.3) | 77.8 |
| 250 – 274 | 37 278 (19.7) | 94.7 | 2 629 (12.0) | 78.8 |
| ≥ 275 | 32 143 (17.0) | 96.0 | 2 126 (9.7) | 83.9 |
| Employment level (%) |  |  |  |  |
| < 65 | 13 396 (7.1) | 89.9 | 6 689 (30.4) | 69.8 |
| 65 – 74 | 43 709 (23.1) | 92.8 | 6 569 (29.9) | 74.8 |
| 75 – 84 | 101 836 (53.8) | 94.1 | 7 270 (33.1) | 79.5 |
| ≥ 85 | 30 211 (16.0) | 95.9 | 1 446 (6.6) | 82.7 |

<sup>a</sup> Postal code area

## Discussion

The observed association between country of birth and vaccine uptake is consistent with a general population survey from the Public Health Agency of Sweden [5]. In this survey, hesitant people expressed concerns about side effects, lacked information, or did not believe they would become seriously ill with COVID-19. Fear of side effects and lack of trust in vaccines have also in international surveys been associated with vaccine hesitancy [6-8]. Furthermore, it is likely that there exist specific barriers for parts of the 70+ population related to e.g. digital identification tools and technical skills required to book time slots for vaccination on-line, which has been standard practice for persons living independently during the Swedish vaccination program.

Another noteworthy finding was the inverse associations between indicators of neighbourhood deprivation and vaccine uptake. This is consistent with results from international general population surveys, reporting more widespread vaccine hesitancy in groups with lower education and income [8, 9]. However, the vaccine uptake was in our study generally satisfactory among native Swedes also in areas with lower socioeconomic status, but dropped substantially among non-Nordic born in those areas. It is conceivable that not only technical barriers but also language and cultural barriers are more prominent in the older population born abroad, but this needs to be explored in detail. The specific reasons for vaccine hesitancy and decline is likely to be heterogeneous, which also calls for further studies.

Our study was limited by the short follow up, why we could not study full vaccine uptake with two doses in relation to the sociodemographic dimensions. However, available data suggested high compliance after the first dose and with no differences in relation to country of birth, which is most likely explained by the standard practice within the program to assist with the scheduling of the second dose in connection with the first visit. The Swedish vaccination program started with the oldest population, why vaccine uptake in the working-age population was too low to permit detailed analysis at the time of this investigation.

In conclusion, the identified clusters of unvaccinated older people, mainly representing ethnic minorities in disadvantaged areas [10], is a public health concern that warrants intensified efforts regarding tailored communication, easier vaccine access and engagement from local communities [2, 7]. Continuous monitoring of the vaccine uptake and immunity of the population is essential for pandemic control and for assessing risks for local or more general disease outbreaks.

## Data Availability

The dataset used in the present study is hosted by the Lund University Population Research Platform (LUPOP). Legal and ethical restrictions prevent public sharing of the dataset. Data can be made available for collaborations upon request to interested researchers but would generally require a new ethical permission. You can find contact information for the data host at https://www.lupop.lu.se/

## Acknowledgments

Cecilia Åkesson-Kotsaris, Paul Söderholm and Helena Hallefjord, Clinical Studies Sweden, for excellence in bringing the data infrastructure in place. Susann Ullén, Clinical Studies Sweden, for statistical advice.

## Author contribution

All authors conceived and designed the study, analyzed and interpreted the results. UM, JB and MM acquired data, JB and MM conducted the statistical analyses. MI and JB drafted the manuscript. All authors critically revised the manuscript and approved the final version for submission.

## Ethics and Permissions

Ethical approval was obtained from the Swedish Ethical Review Authority (2021-00059).

