## Supplementary file for "COVID-19 vaccine uptake among older people in relation to sociodemographic factors –cohort results from southern Sweden"

**Table S1.** COVID-19 vaccine uptake of the study population (n = 212 299 aged 70+) after 160 days of vaccinations, in relation to a detailed stratification of country of birth.

**Table S2.** Multivariable logistic regression model for the association between sociodemographic factors and vaccine uptake in the 70+ population, stratified by country of birth

**Figure S1.** Time to **A)** first dose (n = 216 243 at baseline), **B)** from first to second dose (among persons who received vaccine from Pfizer-BioNTech or Moderna as first dose; n = 147 955), stratified by country of birth (solid curve for Nordic country, dotted curve for non-Nordic Country)

**Table S1.** COVID-19 vaccine uptake of the study population (n = 211 126 aged 70+) after 160 days of vaccinations, in relation to a detailed stratification of country of birth^a^.

| **Country group** | **Included countries** | **N** | **Uptake, %** |
| --- | --- | --- | --- |
| Sweden | Sweden | 181 313 | 94.0 |
| Other Nordic country | Denmark, Norway, Finland, Iceland | 7 839 | 88.9 |
| Western Europe, North America, Australia and New Zeeland | Andorra, Australia, Austria, Belgium, Canada, Cyprus, France, Germany, Greece, Ireland, Italy, Liechtenstein, Luxemburg, Malta, Monaco, Netherlands, New Zeeland, Portugal, San Marino, Spain, Switzerland, UK, USA | 4 542 | 86.3 |
| Eastern Europe, former Yugoslavia, Russia and former republics of Soviet Union | Albania, Bulgaria, Czech republic, Hungary, Poland, Romania, Russia, Slovakia, Turkey, former German Democratic Republic (Eastern Germany) former Yugoslavia, former republics of Soviet union (including the Baltic countries) | 11 974 | 73.0 |
| Middle East | Afghanistan, Bahrain, Iran, Iraq, Israel, Jordan, Lebanon, Palestine, Syria, United Arab Emirates, Yemen | 2 732 | 67.6 |
| Asia, Central and Latin America |  | 1 919 | 76.9 |
| Africa |  | 635 | 66.6 |

^a^Persons born outside the Nordic countries but with unknown country of birth (n = 172) were excluded from this table.

**Table S2.** Multivariable logistic regression model for the association between sociodemographic factors and vaccine uptake, stratified by country of birth.

|  | **All** | **Country of birth** | |
| --- | --- | --- | --- |
|  |  | **Nordic country** | **Other** |
|  | **OR (95% CI)^a^** | **OR (95% CI)^a^** | **OR (95% CI)^a^** |
| **Intercept** | 8.51 (7.85 – 9.23) | 40.2 (36.9 – 43.8) | 6.25 (5.26 – 7.42) |
| **Individual-level characteristics** |  |  |  |
| Born in Nordic country | 4.33 (4.16 – 4.51) | - | - |
| Female | 0.95 (0.92 – 0.99) | 0.97 (0.93 – 1.01) | 0.89 (0.83 – 0.95) |
| Age  70-79  80-89  90+ | 1.00 (Ref.)  1.21 (1.16 -1.26)  1.03 (0.96 – 1.10) | 1.00 (Ref.)  1.17 (1.12-1.22)  1.01 (0.94 – 1.09) | 1.00 (Ref.)  1.31 (1.21 – 1.40)  1.02 (0.88 – 1.18) |
| Civil status  Married  Widow/widower  Divorced  Single | 1.00 (Ref.)  0.73 (0.70 – 0.77)  0.44 (0.42 – 0.46)  0.32 (0.30 – 0.33) | 1.00 (Ref.)  0.69 (0.65 – 0.73)  0.38 (0.37 – 0.40)  0.28 (0.26 – 0.29) | 1.00 (Ref.)  0.85 (0.78 – 0.93)  0.62 (0.58 – 0.67)  0.61 (0.53 – 0.70) |
| **Area-level^b^ characteristics** |  |  |  |
| Born abroad (%)  < 20  20 – 39  40 – 59  ≥ 60 | 1.00 (Ref.)  0.93 (0.88 – 0.97)  0.79 (0.73 – 0.85)  0.77 (0.68 – 0.88) | 1.00 (Ref.)  0.96 (0.91 – 1.02)  0.79 (0.71 – 0.86)  0.63 (0.52-0.78) | 1.00 (Ref.)  0.83 (0.75 – 0.92)  0.76 (0.66 – 0.87)  0.78 (0.65 – 0.93) |
| Education, tertiary (%)  ≥ 60  < 25  25 – 39  40 – 59 | 1.00 (Ref.)  0.65 (0.60 – 0.70)  0.76 (0.71 – 0.80)  0.86 (0.81 – 0.91) | 1.00 (Ref.)  0.57 (0.52 – 0.63)  0.69 (0.64 – 0.75)  0.82 (0.76 – 0.87) | 1.00 (Ref.)  0.86 (0.75 – 0.99)  0.94 (0.84 – 1.07)  0.96 (0.86 – 1.07) |
| Tenants (%)  < 10  10 – 49  50 – 74  ≥ 75 | 1.00 (Ref.)  1.10 (1.06 – 1.15)  0.97 (0.92 – 1.02)  0.91 (0.85 – 0.97) | 1.00 (Ref.)  1.15 (1.10 – 1.21)  1.02 (0.95 – 1.09)  1.02 (0.94 – 1.11) | 1.00 (Ref.)  0.95 (0.87 – 1.04)  0.85 (0.76 – 0.94)  0.72 (0.65 – 0.81) |
| Yearly income, median (kSEK)  ≥ 275  < 200  200 – 249  250 – 274 | 1.00 (Ref.)  0.87 (0.79 – 0.96)  0.89 (0.83 – 0.96)  0.88 (0.82 – 0.94) | 1.00 (Ref.)  0.94 (0.84 – 1.05)  0.93 (0.85 – 1.01)  0.91 (0.84 – 0.99) | 1.00 (Ref.)  0.76 (0.62 – 0.92)  0.84 (0.72 – 1.00)  0.79 (0.68 – 0.93) |
| Employment level (%)  ≥ 85  < 65  65 – 74  75 – 84 | 1.00 (Ref.)  0.77 (0.69 – 0.86)  0.81 (0.75 – 0.88)  0.82 (0.77 – 0.87) | 1.00 (Ref.)  0.73 (0.64 – 0.83)  0.77 (0.70 – 0.85)  0.79 (0.74 – 0.85) | 1.00 (Ref.)  1.03 (0.82 – 1.31)  1.04 (0.85 – 1.27)  1.01 (0.86 – 1.19) |

^a^ Odds ratio (95% confidence interval)

^b^ Postal code area

**Figure S1.** Time to **A)** first dose (n = 216 243 at baseline), **B)** from first to second dose (among persons who received vaccine from Pfizer-BioNTech or Moderna as first dose; n = 147 955), stratified by country of birth (solid curve for Nordic country, dotted curve for non-Nordic Country)

**A**

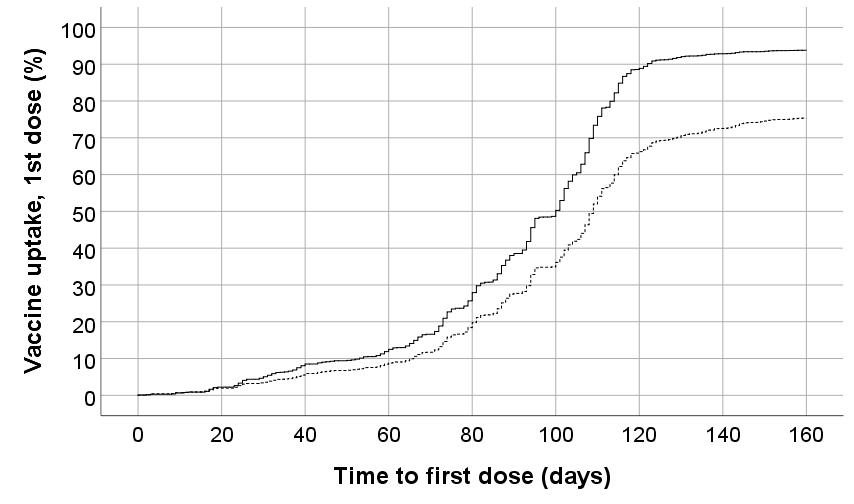

**B**

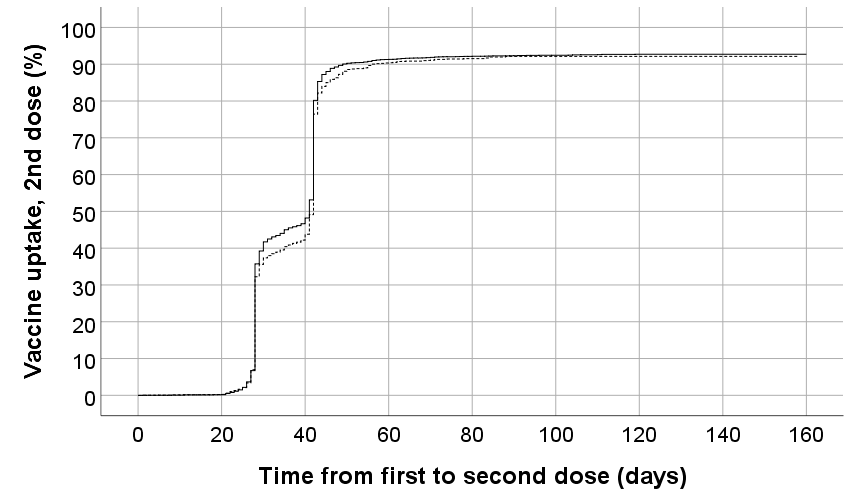
